# Genomic variations of SARS-CoV-2 suggest multiple outbreak sources of transmission

**DOI:** 10.1101/2020.02.25.20027953

**Authors:** Liangsheng Zhang, Jian-Rong Yang, Zhenguo Zhang, Zhenguo Lin

**Affiliations:** College of Agriculture and Biotechnology, Zhejiang University, Hangzhou, China; Zhongshan School of Medicine, Sun Yat-sen University, Guangzhou, China; Independent Scholar, Irvine, CA, 92612. USA; Department of Biology, Saint Louis University, St. Louis, Missouri, USA

## Abstract

We examined 169 genomes of SARS-CoV-2 and found that they can be classified into two major genotypes, Type I and Type II. Type I can be further divided into Type IA and IB. Our phylogenetic analysis showed that the Type IA resembles the ancestral SARS-CoV-2 most. Type II was likely evolved from Type I and predominant in the infections. Our results suggest that Type II SARS-CoV-2 was the source of the outbreak in the Wuhan Huanan market and it was likely originated from a super-spreader. The outbreak caused by the Type I virus should have occurred somewhere else, because the patients had no direct link to the market. Furthermore, by analyzing three genomic sites that distinguish Type I and Type II strains, we found that synonymous changes at two of the three sites confer higher protein translational efficiencies in Type II strains than in Type I strains, which might explain why Type II strains are predominant, implying that Type II is more contagious (transmissible) than Type I. These findings could be valuable for the current epidemic prevention and control.

## Introduction

The outbreak of the coronavirus disease 2019 (COVID-19), caused by a novel coronavirus named “Severe Acute Respiratory Syndrome CoronaVirus 2 (SARS-CoV-2) “, has now been detected in over 60 nations and lead to more than 80,000 confirmed cases and ∼3,000 deaths (http://2019ncov.chinacdc.cn/2019-nCoV/global.html). Although the spread of SARS-CoV-2 in China seems to be largely contained, the confirmed cases outside China have been rising. Therefore, the risk of global pandemic of SARS-CoV-2 is still growing. Currently, the outbreak sources and the transmission history of SARS-CoV-2 is far from well-understood. Revealing the evolutionary history of different SARS-CoV-2 samples allows us to infer the virus transmission routes and to identify novel mutations associated with the transmissions. Such information will be valuable for vaccine development and disease control.

Our recent study showed that SARS-CoV-2 form a sister group to two SARS-like bat viruses (bat-SL-CoVZC45 and bat-SL-CoVZXC21) [1] that were collected in Zhoushan, Zhejiang Province, China, from 2015 to 2017 [2]. A subsequently released sequence of a bat coronavirus (BatCoV RaTG13) isolated in Yunnan in 2013 was found to be more similar to the sequences of SARS-CoV-2 [3], which is confirmed by our phylogenetic analysis based on whole genome sequences (Figure S1). Therefore, we used BatCoV RaTG13 and the two SARS-like bat viruses (bat-SL-CoVZC45 and bat-SL-CoVZXC21) as an outgroup to study the origin and transmission history of SARS-CoV-2.

## Materials and Methods

The complete genomes of SARS-CoV-2 samples were obtained from GISAID (www.gisaid.org), NCBI and NMDC (http://nmdc.cn/#/nCov/). Alignment of the complete genome sequences and that of BatCoV RaTG13 was carried out by MAFFT [4].

We identified genome variable sites from the sequence alignment using Noisy [5]. Variants that are present in at least two samples were used in our subsequence analysis. Some samples have unusually large number of mutation sites, which could be due to due to sequencing or assembly errors. These sequences were also excluded in our analysis.

We inferred the phylogeny of the SARS-CoV-2 isolates based on the variable sites using the maximum likelihood (ML) method by FastTree [6]. The tRNA Adaptation Index (tAI), an indicator of codon translational efficiency, was computed using the tool Bio::CUA (https://metacpan.org/release/Bio-CUA), and the numbers of human tRNA genes for each codon (used for computing tAI) were downloaded from http://gtrnadb.ucsc.edu.

## Results and discussions

We obtained 169 complete genomes of SARS-CoV-2 samples and identified variable sites based on whole genome alignment (Figure 1). In most cases, different SARS-CoV-2 genomes only differ in 0 to 3 sites. A total number of 207 variable sites were identified (Figure S2). It shows that the amplitude of mutations is small, and their evolution history can be inferred from these variable sites.

**Figure 1.**
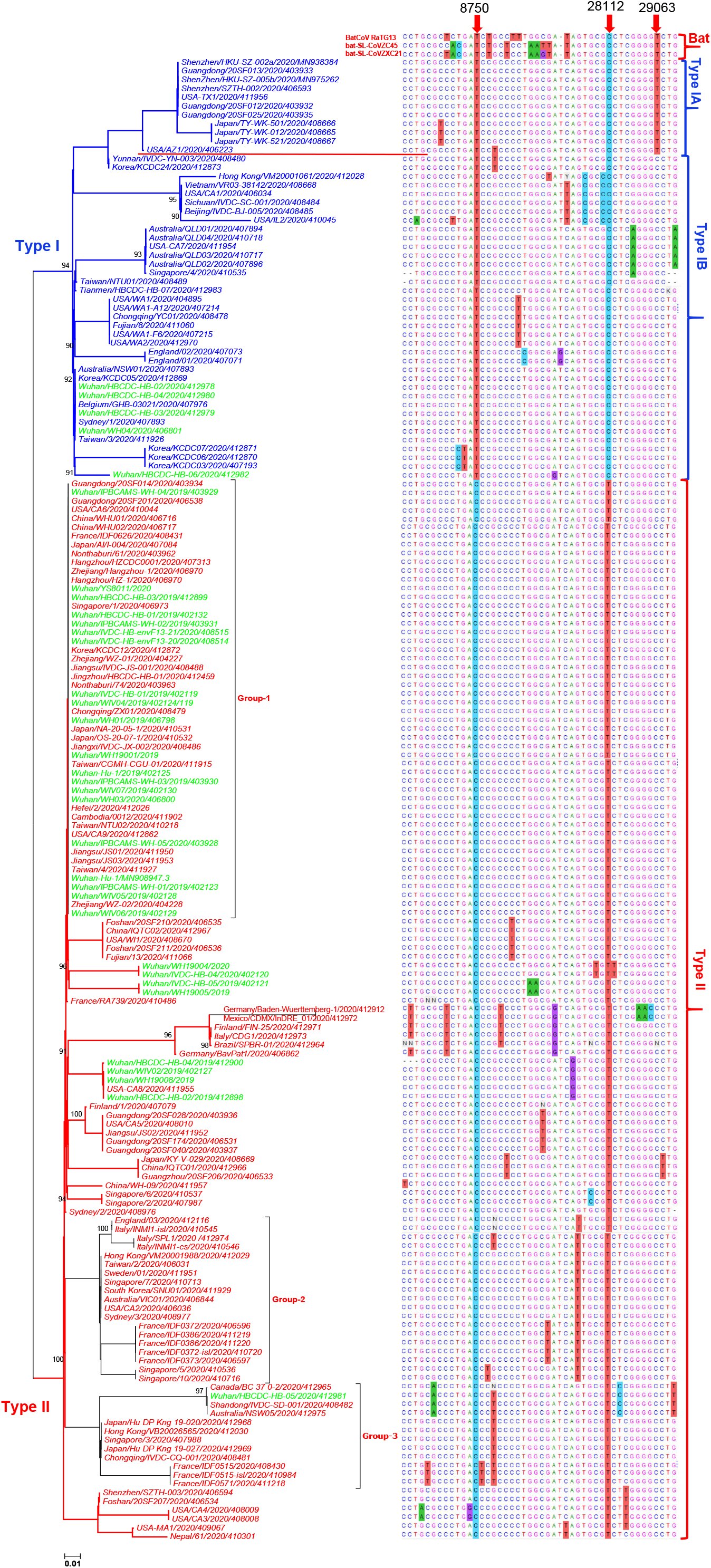
A phylogenetic tree of the 169 SARS-CoV-2strains and their genomic variants. **A**, A maximum likelihood (ML) phylogenetic tree of the human SARS-CoV-2 with approximately ML method by FastTree (http://meta.microbesonline.org/fasttree/). The phylogenetic tree was constructed using the sequence alignment shown in **B**. The two groups, Type I and Type II, are colored in blue and red, respectively. **B**, Sequence alignment of the SARS-CoV-2 genomes where only major variable sites are shown. Each line corresponds to one branch in the phylogenetic tree to the left. The corresponding sites from the strain BatCoV RaTG13 are shown on the top separated by a red line. Three type-specific variants are marked in red arrows, corresponding to the genomic positions 8750, 28112, and 29063, respectively; the coordinates are referred to the sequence MN938384.1.

Their phylogenetic relationships reveal two major genotypes from the SARS-CoV-2 samples, namely Type I and II (Figures 1A, S3 and S4). The genomes of the two types mainly differ at three sites (Figure 1B), which are 8750, 28112, and 29063, using the genome coordinates of sample MN938384.1 as a reference. Specifically, the nucleotides at the three sites are T, C, and T/C in Type I, and C, T, and C in Type II, respectively. Based on the nucleotide at the site 29063, the Type I strains can be further divided into Type IA and IB. The number of genomes belonging to Type IA, IB and II are 11, 37, and 121, respectively.

We found that the three sites in Type IA and two in Type IB are identical to those in three bat viruses BatCoV RaTG13 [3], bat-SL-CoVZC45 and bat-SL-CoVZXC21[2](Figure 1B), suggesting that the Type I may be more closely related to the ancestral human-infecting strain than Type II. Therefore, in the principle of parsimony, Type IA resembles the most ancestral lineage of SARS-CoV-2, and Type IB was derived from Type IA by a new mutation at site 29063. In addition, Type II may have originated from a Type IB strain by accumulating mutations at sites 8750 and 28112.

Two environmental samples isolated from the Huanan market (Wuhan/IVDC-HB-envF13-20 and 21) belong to Type II, and no samples from Type I have a direct link to the Huanan market (such as Wuhan/WH04/2020 [7]). These observations suggest that the outbreak in the Huanan market was triggered by the Type II virus and that the initial transmission of Type I viruses to humans might have occurred somewhere else in Wuhan, probably preceding the outbreak of Type II in the Huanan market. Our speculation is in line with earlier reports that 14 of the first reported 41 cases had no link to the Huanan market [7-9].

In addition to the above-mentioned transmission sources, we also found that there are several large clades in Type II (Group 1-3). The Group-1 of Type II are identical, suggesting they were likely originated from the same transmission source. Group-2 and Group-3 share at least one mutation, indicating that they shared the same transmission source. As only a small portion of SARS-CoV-2 isolates have been sequenced, the actual numbers of infected patients in each group are expected to be much larger. We speculate that each group was likely from a source of super transmission or super-spreader.

Because Type II strains account for most of the sampled genomes, it is reasonable to speculate that Type II is more contagious, assuming no strong sampling bias. To better understand why the Type II virus is more prevalent, we then focused on the three variable genomic sites that distinguish Type II from Type I strains. First, we noticed the mutations at 8750 and 29063 are synonymous (in gene *orf1ab* and *N*, respectively) and the one at 28112 is nonsynonymous, leading to a change from Leucine to Serine in the gene *ORF8*. Interestingly, we found that the two synonymous changes both confer higher translational efficiencies for the Type II strains than for Type I (Figure 2), based on the number of tRNA genes matching each codon and tRNA Adaptation Index (tAI) [10]. We speculate that the higher translational efficiencies might have enabled faster production of Type II virus particles, facilitated its spread, and led to its dominance in infections, implying that Type II is more contagious (transmissible) than Type I. If our speculation is correct, this would provide guidelines for treating Type I and II patients differently.

**Figure 2.**
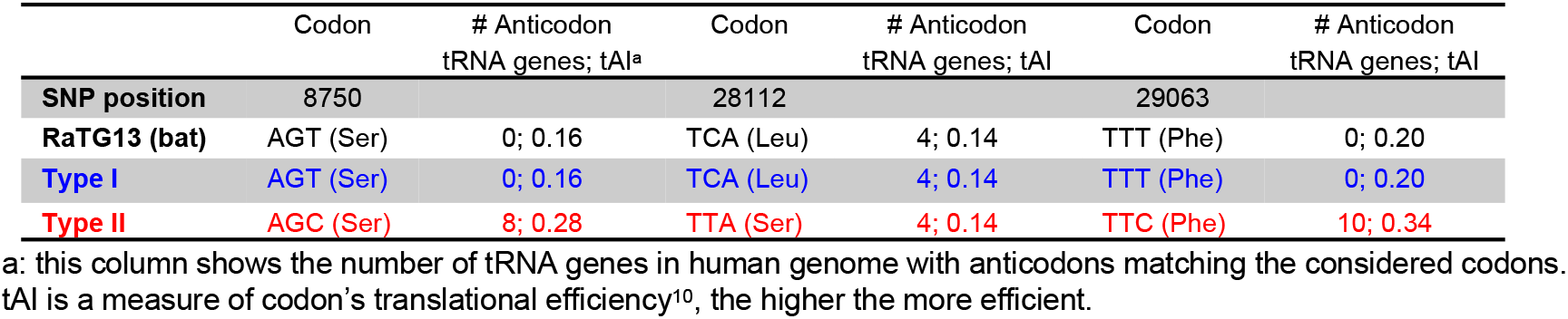
The codon changes caused by the three variant sites in Figure 1. The tAI values were computed using Bio::CUA (https://metacpan.org/release/Bio-CUA), and the numbers of human tRNA genes were downloaded from http://gtrnadb.ucsc.edu.

In summary, our results illustrate the presence of two major genotypes of SARS-CoV-2, suggesting of at least two, possibly three, major outbreak sources (Figure 3). The outbreak in the Huanan market may not be the initiation transmission of SARS-CoV-2 to human, and thus the location of initial transmission to humans remain to be determined. As most samples detected belong to Type II, it indicates that Type II is more transmissible. We identified three genomic variants that separate the two types of SARS-CoV-2. Our data suggest that two variants are linked to improved translation efficiency in Type II. The impacts of these genomic variants could be further evaluated by comparing the symptoms of patients infected by each type of viruses. Particularly, some asymptomatic carriers have been recently found [11]. It is worth examining whether these asymptomatic cases are enriched in Type I. With more sequencing data of SARS-CoV-2, we expect a more complete understanding of transmission history.

**Figure 3.**
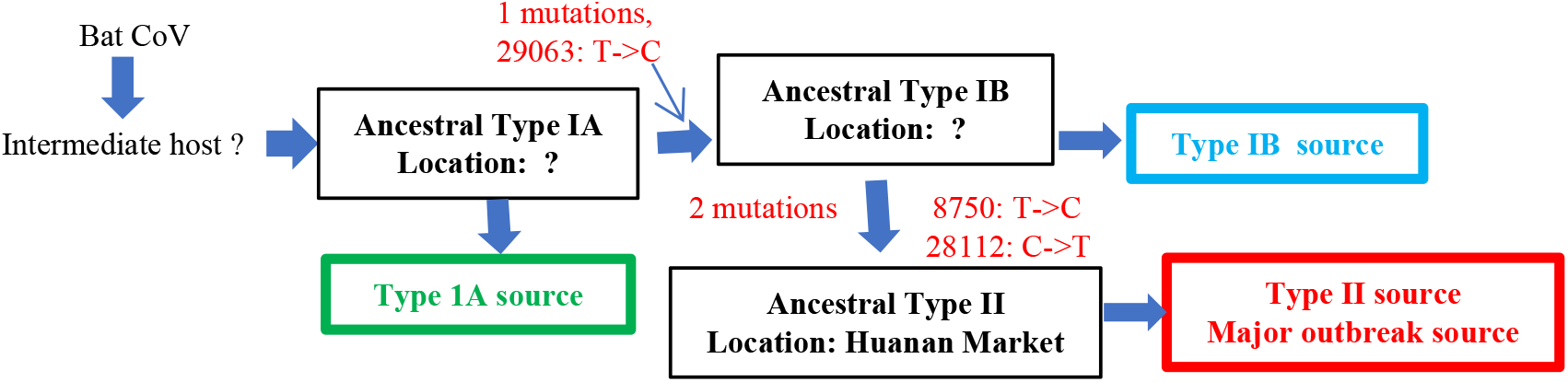
A simple SARS-CoV-2 virus transmission model. The SARS-CoV-2 may have two or three sources of transmissions, corresponding to the three genotypes we identified here, Type IA, Type IB and Type II.

## Data Availability

We obtained 97 complete genomes of COVID-19 samples from GISAID (www.gisaid.org), NCBI and NMDC (http://nmdc.cn/#/nCov/).

## Acknowledgments

We acknowledge the authors and the originating and submitting laboratories of the nucleotide sequences from the Global Initiative on Sharing All Influenza Data’s EpiFlu Database, NCBI and NMDC (http://nmdc.cn/#/nCov/).

---

**Supplementary Figure 1**. The SARS-CoV and SARS-CoV-2 phylogenetic tree uses MERS-CoV as an outgroup.

**Supplementary Figure 2**. Sequence alignment of 169 COVID-19 genomes where only the variable sites are shown.

**Supplementary Figure 3**. A maximum likelihood (ML) phylogenetic tree of the human COVID-19 virus. The phylogenetic tree was constructed using the genome sequence alignment. The two groups, Type I and Type II, are colored in blue and red, respectively.

**Supplementary Figure 4**. A maximum likelihood (ML) phylogenetic tree of the human COVID-19 virus. The phylogenetic tree was constructed using the variable sites shown in **Supplementary Figure 2**. The two groups, Type I and Type II, are colored in blue and red, respectively.

## References

1. Zhang L., et al., Origin and evolution of the 2019 novel coronavirus. Clin Infect Dis, 2020.

2. Hu, D., et al., Genomic characterization and infectivity of a novel SARS-like coronavirus in Chinese bats. Emerg Microbes Infect, 2018. 7(1): p. 154.

3. Zhou, jP., et al., A pneumonia outbreak associated with a new coronavirus of probable bat origin. Nature, 2020.

4. Yamada, K.D., K. Tomii, and K. Katoh, Application of the MAFFT sequence alignment program to large data-reexamination of the usefulness of chained guide trees. Bioinformatics, 2016. 32(3): p. 3246–3251.

5. Dress, A.W., et al., Noisy: identification of problematic columns in multiple sequence alignments. Algorithms Mol Biol, 2008. 3: p. 7.

6. Price, M.N., P.S. Dehal, and A.P. Arkin, FastTree 2--approximately maximum-likelihood trees for large alignments. PLoS One, 2010. 5(i3): p. e9490.

7. Lu, R., et al., Genomic characterisation and epidemiology of 2019 novel coronavirus: implications for virus origins and receptor binding. The Lancet.

8. Huang, C., et al., Clinical features of patients infected with 2019 novel coronavirus in Wuhan, China. Lancet, 2020.

9. Li, Q., et al., Early Transmission Dynamics in Wuhan, China, of Novel Coronavirus–Infected Pneumonia. New England Journal of Medicine, 2020.

10. dos Reis, M., R. Savva, and L. Wernisch, Solving the riddle of codon usage preferences: a test for translational selection. Nucleic Acids Res, 2004. 32(17): p. 5036–44.

11. Bai, Y., et al., Presumed Asymptomatic Carrier Transmission of COVID-19. JAMA, 2020.

